# Heterogeneity and temporal variation in the management of COVID-19: a multinational drug utilization study including 71,921 hospitalized patients from China, South Korea, Spain, and the United States of America

**DOI:** 10.1101/2020.09.15.20195545

**Authors:** Albert Prats-Uribe, Anthony G. Sena, Lana Yin Hui Lai, Waheed-Ul-Rahman Ahmed, Heba Alghoul, Osaid Alser, Thamir M Alshammari, Carlos Areia, William Carter, Paula Casajust, Dalia Dawoud, Asieh Golozar, Jitendra Jonnagaddala, Paras P. Mehta, Mengchun Gong, Daniel R. Morales, Fredrik Nyberg, Jose D. Posada, Martina Recalde, Elena Roel, Karishma Shah, Nigam H. Shah, Lisa M. Schilling, Vignesh Subbian, David Vizcaya, Andrew Williams, Lin Zhang, Ying Zhang, Hong Zhu, Li Liu, Peter Rijnbeek, George Hripcsak, Jennifer CE Lane, Edward Burn, Christian Reich, Marc A. Suchard, Talita Duarte-Salles, Kristin Kostka, Patrick Ryan, Daniel Prieto-Alhambra

**Author notes:** Corresponding author: Patrick Ryan, Janssen Research & Development, Titusville, NJ, USA. Joint first authors (Albert Prats-Uribe, Anthony G. Sena).

## Abstract

**Objectives:** A plethora of medicines have been repurposed or used as adjunctive therapies for COVID-19. We characterized the utilization of medicines as prescribed in routine practice amongst patients hospitalized for COVID-19 in South Korea, China, Spain, and the USA.

**Design:** International network cohort

**Setting:** Hospital electronic health records from Columbia University Irving Medical Centre (NYC, USA), Stanford (CA, USA), Tufts (MA, USA), Premier (USA), Optum EHR (USA), department of veterans affairs (USA), NFHCRD (Honghu, China) and HM Hospitals (Spain); and nationwide claims from HIRA (South Korea)

**Participants:** patients hospitalized for COVID-19 from January to June 2020

**Main outcome measures:** Prescription/dispensation of any medicine on or 30 days after hospital admission date

**Analyses:** Number and percentage of users overall and over time

**Results:** 71,921 people were included: 304 from China, 2,089 from Spain, 7,599 from South Korea, and 61,929 from the USA. A total of 3,455 medicines were identified. Common repurposed medicines included hydroxychloroquine (<2% in NFHCRD to 85.4% in HM), azithromycin (4.9% in NFHCRD to 56.5% in HM), lopinavir/ritonavir (<3% in all US but 34.9% in HIRA and 56.5% in HM), and umifenovir (0% in all except 78.3% in NFHCRD). Adjunctive medicines were used with great variability, with the ten most used treatments being (in descending order): bemiparin, enoxaparin, heparin, ceftriaxone, aspirin, vitamin D, famotidine, vitamin C, dexamethasone, and metformin. Hydroxychloroquine and azithromycin increased rapidly in use in March-April but declined steeply in May-June.

**Conclusions:** Multiple medicines were used in the first months of COVID-19 pandemic, with substantial geographic and temporal variation. Hydroxychloroquine, azithromycin, lopinavir-ritonavir, and umifenovir (in China only) were the most prescribed repurposed medicines. Antithrombotics, antibiotics, H2 receptor antagonists and corticosteroids were often used as adjunctive treatments. Research is needed on the comparative risk and benefit of these treatments in the management of COVID-19.

What is already known in this topic

- Drug repurposing is a common approach in the clinical management of novel diseases and conditions for which there are no available pharmacotherapies
- Hydroxychloroquine was widely used in the management of COVID-19 patients during the early phases of the pandemic
- Recent NIH (and other) guidelines recommend the use of concomitant therapies including immune-based, antithrombotic, antibiotic and other treatments

What this study adds

- This study demonstrates great variability and extensive drug repurposing and utilization in the management of COVID-19 patients.
- A wide range of adjunctive treatments has been used, including antithrombotics, antibiotics, H2 receptor antagonists, and systemic corticosteroids.
- Emerging clinical data on the safety and efficacy of hydroxychloroquine and azithromycin impacted their rise and rapid decline in use internationally
- Conversely, the use of corticosteroids grew only in more recent months, with little use in the early stages of the pandemic (January to April)

## Background

As of September 01, 2020, there were more than 25 million confirmed cases of coronavirus disease 2019 (COVID-19) and more than 850,000 related deaths worldwide ^1^. Despite a lack of evidence, several medicines were repurposed in the first months of the pandemic based on *in vitro* anti-viral activity. As an illustrative example, hydroxychloroquine obtained emergency approval by the US Food and Drug Administration on March 28, 2020 which was later revoked on June 15, 2020^th 2^. More recently, a few investigational drugs have been tested, with remdesivir being the frontrunner antiviral after an international placebo-controlled randomized controlled trial (RCT) showed promising results ^3^.

In the absence of approved antivirals, the cornerstone of management has been supportive care, where adjunctive therapies play a major role. Three recognized adjunctive therapies in COVID-19 are corticosteroids, anti-cytokines (e.g., tocilizumab), and immunoglobulins (e.g. convalescent plasma) ^4^. Of these, dexamethasone, and corticosteroids, have recently been shown to reduce mortality among patients receiving mechanical ventilation or oxygen therapy in a large RCT ^5,6^. Additional adjunctive therapies are recognised in recent guidelines, including antithrombotics, statins, antihypertensives and other concomitant treatments. Clinical guidelines have varied in their recommendations on COVID-19 treatment geographically and over time ^7^. In these circumstances, it is crucial to understand the use of treatments, identify trends of prescribing and determine rational medication use in different health care settings.

We aimed to characterize the use of repurposed and adjunctive medicines among patients hospitalized for COVID-19, and amongst those receiving intensive care in actual practice settings across Europe, Asia, and North America.

## Methods

### Study Design

Multinational network cohort study based on hospital electronic health records (EHRs), and claims data. Data from different sites were mapped to the Observational Medical Outcomes Partnership (OMOP) Common Data Model (CDM) ^8^. This approach allowed contributing centers to execute analytical code in a distributed/federated fashion without sharing patient-level data.

### Data Sources

Data were obtained from USA, South Korea, China and Spain. EHR data from the USA came from Columbia University Irving Medical Center (CUIMC), STAnford medicine Research data Repository (STARR-OMOP)^9^, Premier, Optum© de-identified Electronic Health Record Dataset (OPTUM), Tufts CLARET (TRDW), and the Department of Veterans Affairs (VA). Data from South Korea came from nation-wide claims recorded in the Health Insurance Review & Assessment (HIRA). Inpatient EHR data from Spain was obtained from HM Hospitals (HM). Data from China was extracted from nine hospitals in Honghu, supported by Nanfang Hospital, Southern Medical University, and contained full EMR data (NFHCRD). Intensive care drug use data was available from Premier, OPTUM, and VA-OMOP. A detailed description of the databases can be found in Appendix Table 1.

**Table 1.**
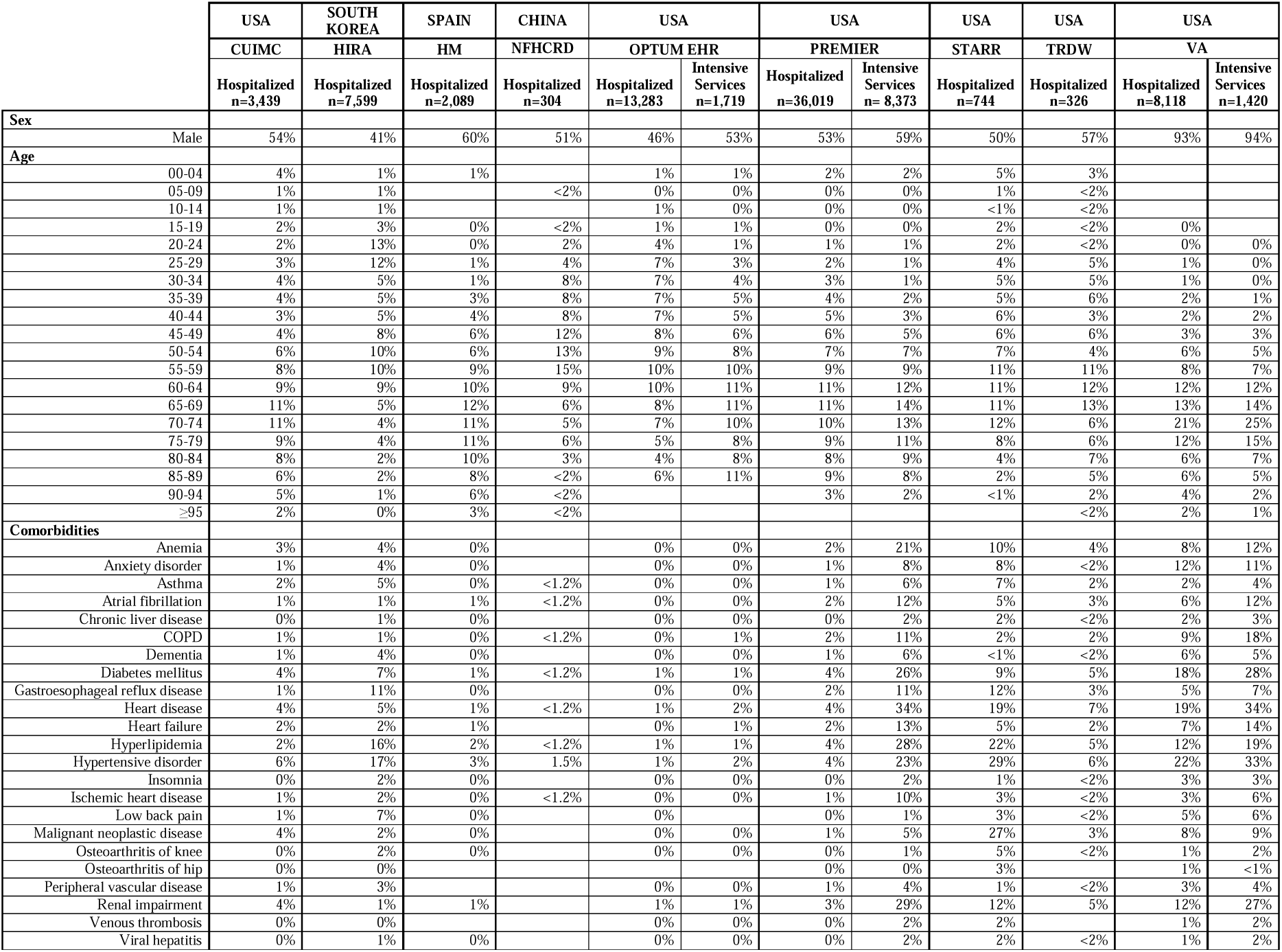
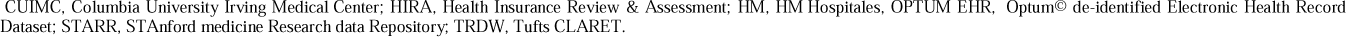
Baseline Characteristics of study participants, stratified by data source.

|  |  | USA | SOUTH KOREA | SPAIN | CHINA | USA |  | USA |  | USA | USA | USA |  |
| --- | --- | --- | --- | --- | --- | --- | --- | --- | --- | --- | --- | --- | --- |
|  |  | CUIMC | HIRA | HM | NFHC RD | OPTUM EHR |  | PREMIER |  | STARR | TRDW | VA |  |
|  |  | Hospitalized<br>n=3,439 | Hospitalized<br>n=7,599 | Hospitalized<br>n=2,089 | Hospitalized<br>n=304 | Hospitalized<br>n=13,283 | Intensive<br>Services<br>n=1,719 | Hospitalized<br>n=36,019 | Intensive<br>Services<br>n= 8,373 | Hospitalized<br>n=744 | Hospitalized<br>n=326 | Hospitalized<br>n=8,118 | Intensive<br>Services<br>n=1,420 |
| **Sex** |  |  |  |  |  |  |  |  |  |  |  |  |  |
|  | Male | 54% | 41% | 60% | 51% | 46% | 53% | 53% | 59% | 50% | 57% | 93% | 94% |
| **Age** |  |  |  |  |  |  |  |  |  |  |  |  |  |
|  | 00-04 | 4% | 1% | 1% |  | 1% | 1% | 2% | 2% | 5% | 3% |  |  |
|  | 05-09 | 1% | 1% |  | <2% | 0% | 0% | 0% | 0% | 1% | <2% |  |  |
|  | 10-14 | 1% | 1% |  |  | 1% | 0% | 0% | 0% | <1% | <2% |  |  |
|  | 15-19 | 2% | 3% | 0% | <2% | 1% | 1% | 0% | 0% | 2% | <2% | 0% |  |
|  | 20-24 | 2% | 13% | 0% | 2% | 4% | 1% | 1% | 1% | 2% | <2% | 0% | 0% |
|  | 25-29 | 3% | 12% | 1% | 4% | 7% | 3% | 2% | 1% | 4% | 5% | 1% | 0% |
|  | 30-34 | 4% | 5% | 1% | 8% | 7% | 4% | 3% | 1% | 5% | 5% | 1% | 0% |
|  | 35-39 | 4% | 5% | 3% | 8% | 7% | 5% | 4% | 2% | 5% | 6% | 2% | 1% |
|  | 40-44 | 3% | 5% | 4% | 8% | 7% | 5% | 5% | 3% | 6% | 3% | 2% | 2% |
|  | 45-49 | 4% | 8% | 6% | 12% | 8% | 6% | 6% | 5% | 6% | 6% | 3% | 3% |
|  | 50-54 | 6% | 10% | 6% | 13% | 9% | 8% | 7% | 7% | 7% | 4% | 6% | 5% |
|  | 55-59 | 8% | 10% | 9% | 15% | 10% | 10% | 9% | 9% | 11% | 11% | 8% | 7% |
|  | 60-64 | 9% | 9% | 10% | 9% | 10% | 11% | 11% | 12% | 11% | 12% | 12% | 12% |
|  | 65-69 | 11% | 5% | 12% | 6% | 8% | 11% | 11% | 14% | 11% | 13% | 13% | 14% |
|  | 70-74 | 11% | 4% | 11% | 5% | 7% | 10% | 10% | 13% | 12% | 6% | 21% | 25% |
|  | 75-79 | 9% | 4% | 11% | 6% | 5% | 8% | 9% | 11% | 8% | 6% | 12% | 15% |
|  | 80-84 | 8% | 2% | 10% | 3% | 4% | 8% | 8% | 9% | 4% | 7% | 6% | 7% |
|  | 85-89 | 6% | 2% | 8% | <2% | 6% | 11% | 9% | 8% | 2% | 5% | 6% | 5% |
|  | 90-94 | 5% | 1% | 6% | <2% |  |  | 3% | 2% | <1% | 2% | 4% | 2% |
|  | ≥95 | 2% | 0% | 3% | <2% |  |  |  |  |  | <2% | 2% | 1% |
| **Comorbidities** |  |  |  |  |  |  |  |  |  |  |  |  |  |
|  | Anemia | 3% | 4% | 0% |  | 0% | 0% | 2% | 21% | 10% | 4% | 8% | 12% |
|  | Anxiety disorder | 1% | 4% | 0% |  | 0% | 0% | 1% | 8% | 8% | <2% | 12% | 11% |
|  | Asthma | 2% | 5% | 0% | <1.2% | 0% | 0% | 1% | 6% | 7% | 2% | 2% | 4% |
|  | Atrial fibrillation | 1% | 1% | 1% | <1.2% | 0% | 0% | 2% | 12% | 5% | 3% | 6% | 12% |
|  | Chronic liver disease | 0% | 1% | 0% |  | 0% | 0% | 0% | 2% | 2% | <2% | 2% | 3% |
|  | COPD | 1% | 1% | 0% | <1.2% | 0% | 1% | 2% | 11% | 2% | 2% | 9% | 18% |
|  | Dementia | 1% | 4% | 0% |  | 0% | 0% | 1% | 6% | <1% | <2% | 6% | 5% |
|  | Diabetes mellitus | 4% | 7% | 1% | <1.2% | 1% | 1% | 4% | 26% | 9% | 5% | 18% | 28% |
|  | Gastroesophageal reflux disease | 1% | 11% | 0% |  | 0% | 0% | 2% | 11% | 12% | 3% | 5% | 7% |
|  | Heart disease | 4% | 5% | 1% | <1.2% | 1% | 2% | 4% | 34% | 19% | 7% | 19% | 34% |
|  | Heart failure | 2% | 2% | 1% |  | 0% | 1% | 2% | 13% | 5% | 2% | 7% | 14% |
|  | Hyperlipidemia | 2% | 16% | 2% | <1.2% | 1% | 1% | 4% | 28% | 22% | 5% | 12% | 19% |
|  | Hypertensive disorder | 6% | 17% | 3% | 1.5% | 1% | 2% | 4% | 23% | 29% | 6% | 22% | 33% |
|  | Insomnia | 0% | 2% | 0% |  | 0% | 0% | 0% | 2% | 1% | <2% | 3% | 3% |
|  | Ischemic heart disease | 1% | 2% | 0% | <1.2% | 0% | 0% | 1% | 10% | 3% | <2% | 3% | 6% |
|  | Low back pain | 1% | 7% | 0% |  | 0% |  | 0% | 1% | 3% | <2% | 5% | 6% |
|  | Malignant neoplastic disease | 4% | 2% | 0% |  | 0% | 0% | 1% | 5% | 27% | 3% | 8% | 9% |
|  | Osteoarthritis of knee | 0% | 2% | 0% |  | 0% | 0% | 0% | 1% | 5% | <2% | 1% | 2% |
|  | Osteoarthritis of hip | 0% | 0% |  |  |  |  | 0% | 0% | 3% |  | 1% | <1% |
|  | Peripheral vascular disease | 1% | 3% |  |  | 0% | 0% | 1% | 4% | 1% | <2% | 3% | 4% |
|  | Renal impairment | 4% | 1% | 1% |  | 1% | 1% | 3% | 29% | 12% | 5% | 12% | 27% |
|  | Venous thrombosis | 0% | 0% |  |  | 0% | 0% | 0% | 2% | 2% |  | 1% | 2% |
|  | Viral hepatitis | 0% | 1% | 0% |  | 0% | 0% | 0% | 2% | 2% | <2% | 1% | 2% |

CUIMC, Columbia University Irving Medical Center; HIRA, Health Insurance Review & Assessment; HM, HM Hospitales, OPTUM EHR, Optum© de-identified Electronic Health Record Dataset; STARR, STANford medicine Research data Repository; TRDW, Tufts CLARET.

### Study Participants

Patients hospitalized with a recorded diagnosis of COVID-19 or a positive test result for SARS-CoV-2 between January and June 2020 were included. A second cohort of patients receiving intensive services was identified as a subset of the former, defined by the initiation of mechanical ventilation, extracorporeal membrane oxygenation/ECMO, and/or tracheostomy. Index dates were the date of admission to hospital and the date of start of intensive services respectively.

### Drugs of interest

All medicines prescribed/dispensed during hospital admission and in the month prior were ascertained for characterization. For the study of medicines used for COVID-19 we assessed all medicines included in at least two RCTs according to the COVID-19 clinical trial tracker ^10^. The resulting list was circulated to stakeholders with a role in drug development and research (e.g., key opinion leaders, pharma industry) and drug regulatory agencies, with all suggestions added to the list. Drugs were classified into two groups: i) *repurposed* medicines - those with alternative indications but believed to be efficacious as antivirals, ii) *adjuvant* therapies - drugs used for the treatment of pneumonia or preventing or treating COVID-19 complications ^4^.

### Statistical analyses

Age, sex, and history of medical conditions were summarized as proportions, as calculated by the number of persons within a given category, divided by the total number of persons.

Medication use was calculated over two time periods: (1) from 1 to 30 days before index date; and (2) from index date to 30 days after. Episodes of drug/s use were defined as starting on the date of first drug exposure and ending on the observed end date (if available), or inferred (for example, based on the number of days of supply), with a persistence window of <=30 days permitted between two prescriptions ^11^. We computed prevalence of use for each drug and major drug classes in both time periods. Prevalence of medication use for each time window was determined by the proportion of subjects who had >=1 day during the time window overlapping with a drug use period for each medication or drug class of interest. All drugs and additional time windows (a year prior and on index date) are reported in full and will be updated over time as more data become available in a dedicated interactive website (https://data.ohdsi.org/Covid19CharacterizationCharybdis/). All (aggregated) data can be downloaded from this same website.

As an initial approach to characterize all use of medicines, rainbow plots were generated for each database. These display the proportion of users of each medicine in the days 0 to 30 from index, using Anatomical Therapeutic Chemical (ATC) groupings

To curtail the long list of adjuvant treatments, we compared the prevalence of use of *adjuvant* therapies on -30 to -1 days before diagnosis and 0 to 30 days after (for all databases in which pre-index data was available). For this, we computed standardized mean differences (SMD), a widely used method to detect differences, and selected those with SMD>0.1; or those without use before diagnosis, for display in the main figures.

We created lollipop plots of cumulative incidence of drug use 0 to 30 days after diagnosis and after hospitalization for *repurposed* and for *adjuvant* drugs. Graphs with the whole list of adjuvants drugs can be found in the supplementary material.

We calculated cumulative incidence of drug use 0 to 30 days after diagnosis for the selected drugs by month of index date. To ensure enough time points, we selected databases with 3 or more months of data available. We plotted cumulative incidence of use per calendar month in the study period.

## Results

A total of 10 databases were analyzed, including 71,921 participants: 2,089 from Spain, 7,599 from South Korea, 304 from China, 744 from California, 326 from Massachusetts, 3,493 from New York, 8,118 from US-wide VA, and the remaining 49,302 from US-wide databases (Premier and OptumEHR). Of these, 11,512 participants (from VA, Premier, and OptumEHR) were included in the intensive care cohorts.

All the results from this study are available as an interactive website (https://data.ohdsi.org/Covid19CharacterizationCharybdis/). This website contains both the summary results presented here, and further details including all medications and comorbidities recorded for the cohorts of interest.

Baseline characteristics are detailed in Table 1. Age varied slightly across data sources, but most cases clustered around the ages of 50 to 74 years old. There was a majority of men amongst those admitted in China (51%), Spain (60%) and the US (50% to 94%), but not in South Korea (41%).

A total of 3,455 different medicines were administered to the study participants in the month after hospitalization, as depicted in Figure 1. The ATC groups *anti-infectives for systemic use*, treatments for ‘*blood and blood forming organs*’, ‘*cardiovascular system’* therapies, and drugs for the ‘*musculoskeletal system*’ were consistently seen amongst the most commonly prescribed.

**Figure 1.**
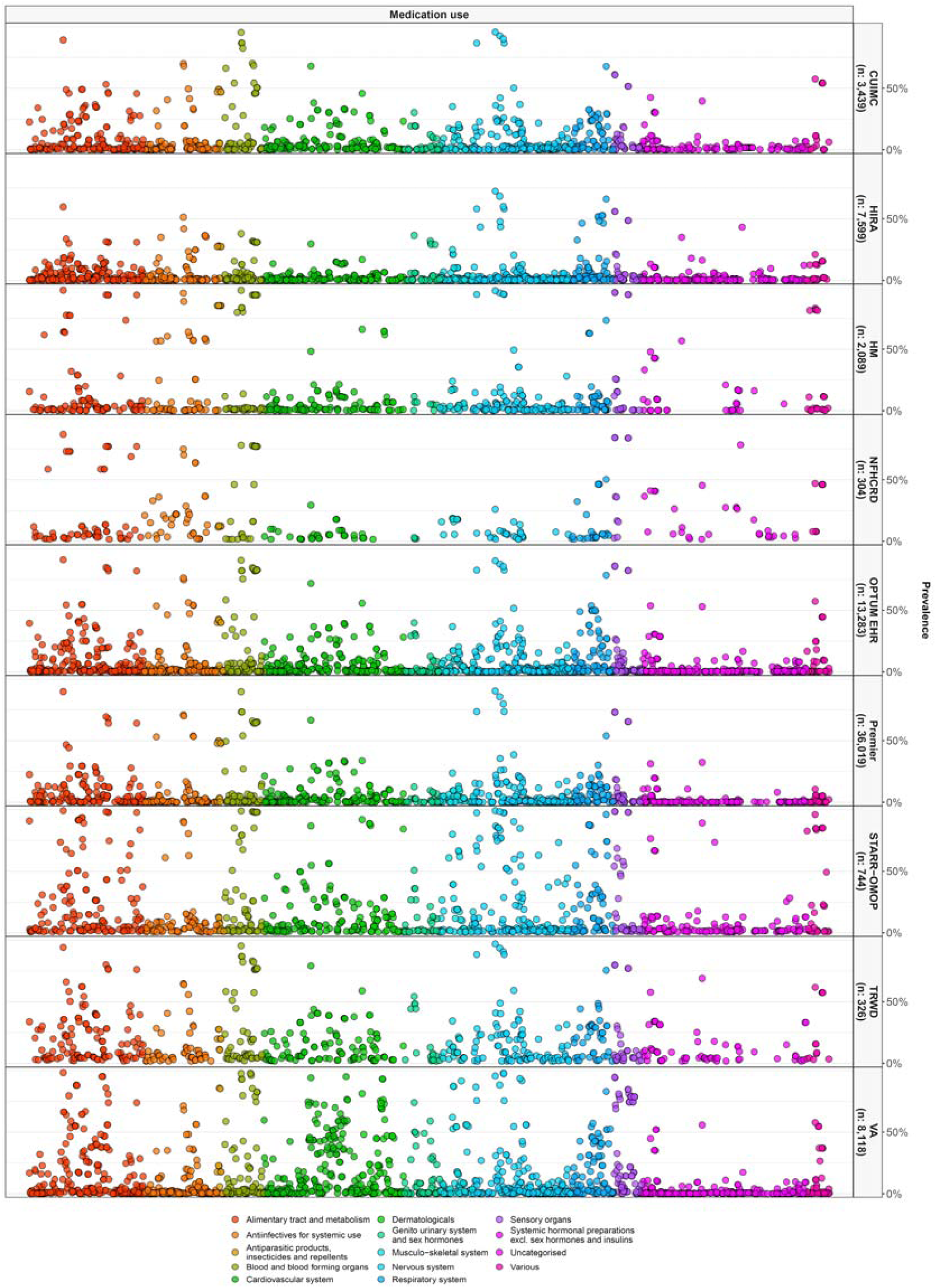
Percentage (%) of 30-day use of all medicines (rainbow plot) in hospitalized patients.

Among the targeted drugs, the top 10 most common in each of the contributing databases are reported in Table 2. In addition to hydroxychloroquine, ritonavir, lopinavir, oseltamivir, remdesivir and umifenovir were the most popular antivirals, with the latter used exclusively in China. Commonly used adjunctive therapies included antibiotics, antithrombotics, corticosteroids, metformin, vitamin (C and D) supplements, antihypertensives, H2 receptor antagonists, and interleukin inhibitors.

**Table 2a.** Top 5 most used *repurposed* drugs in each data source.

|  | CUIMC |  | HIRA |  | HM |  | NFHCRD |  | OPTUM EHR |  |  |  |
| --- | --- | --- | --- | --- | --- | --- | --- | --- | --- | --- | --- | --- |
|  | Hospitalized |  | Hospitalized |  | Hospitalized |  | Hospitalized |  | Hospitalized |  | Intensive Services |  |
| N | Treatment | % | Treatment | % | Treatment | % | Treatment | % | Treatment | % | Treatment | % |
| 1 | Hydroxychloroquine | 46.9% | Lopinavir | 34.9% | Hydroxychloroquine | 85.4% | Umifenovir | 78.3% | Azithromycin | 53.5% | Azithromycin | 46.4% |
| 2 | Azithromycin | 31.7% | Ritonavir | 34.9% | Ritonavir | 56.8% | Ribavirin | 21.1% | Hydroxychloroquine | 40.0% | Hydroxychloroquine | 37.8% |
| 3 | Oseltamivir | 1.7% | Hydroxychloroquine | 27.4% | Lopinavir | 56.8% | Oseltamivir | 13.2% | Lopinavir | 2.8% | Ritonavir | 4.6% |
| 4 | Ivermectin | 0.2% | Azithromycin | 13.7% | Azithromycin | 56.5% | Chloroquine | 11.5% | Ritonavir | 2.8% | Lopinavir | 4.6% |
| 5 | Ritonavir | 0.2% | Peginterferon alfa-2a | 0.4% | Oseltamivir | 6.9% | Lopinavir | 7.2% | Oseltamivir | 0.8% | Oseltamivir | 0.6% |

|  | PREMIER |  |  |  | STARR-OMOP |  | TRDW |  | VA |  |  |  |
| --- | --- | --- | --- | --- | --- | --- | --- | --- | --- | --- | --- | --- |
|  | Hospitalized |  | Intensive Services |  | Hospitalized |  | Hospitalized |  | Hospitalized |  | Hospitalized |  |
| N | Treatment | % | Treatment | Treatment | Treatment | Treatment | Treatment | % | Treatment | % | Treatment | % |
| 1 | Azithromycin | 53.1% | Hydroxychloroquine | Azithromycin | Azithromycin | Azithromycin | Azithromycin | 28.5% | Azithromycin | 47.5% | Azithromycin | 52.2% |
| 2 | Hydroxychloroquine | 47.9% | Azithromycin | Hydroxychloroquine | Hydroxychloroquine | Hydroxychloroquine | Hydroxychloroquine | 19.9% | Hydroxychloroquine | 22.7% | Hydroxychloroquine | 47.8% |
| 3 | Oseltamivir | 1.2% | Oseltamivir | Oseltamivir | Oseltamivir | Oseltamivir | Oseltamivir |  | Remdesivir | 1.8% |  |  |
| 4 | Ritonavir | 0.7% | Ritonavir | Ritonavir | Ritonavir | Ritonavir | Ritonavir |  | Oseltamivir | 1.7% |  |  |
| 5 | Lopinavir | 0.6% | Lopinavir | Ivermectin | Ivermectin | Ivermectin | Ivermectin |  | Ritonavir | 0.5% |  |  |

**Table 2b.** Top 10 most used adjunctive drugs in each data source.

|  | CUIMC |  | HIRA |  | HM |  | NFHCRD |  | OPTUM EHR |  |  |  |
| --- | --- | --- | --- | --- | --- | --- | --- | --- | --- | --- | --- | --- |
|  | Hospitalized |  | Hospitalized |  | Hospitalized |  | Hospitalized |  | Hospitalized |  | Intensive Services |  |
| N | Treatment | % | Treatment | % | Treatment | % | Treatment | % | Treatment | % | Treatment | % |
| 1 | Enoxaparin | 66.2% | Fluoroquinolones | 24.7% | Ceftriaxone | 60.5% | Fluoroquinolones | 63.8% | Enoxaparin | 58.3% | Heparin | 61.3% |
| 2 | Ceftriaxone | 40.6% | H2 receptor antagonist | 16.4% | Corticosteroids | 42.8% | Vitamin C | 58.6% | Ceftriaxone | 46.3% | Enoxaparin | 56.5% |
| 3 | Heparin | 35.1% | Statins | 13.5% | Fluoroquinolones | 25.6% | Corticosteroids | 40.8% | Statins | 38.0% | H2 receptor antagonist | 54.3% |
| 4 | Statins | 32.5% | ARBs | 13.1% | Interleukin inhibitors | 16.4% | Immunoglobulins | 22.0% | Heparin | 34.3% | Famotidine | 54.2% |
| 5 | Corticosteroids | 30.4% | Famotidine | 10.5% | Tocilizumab | 16.3% | Amoxicillin | 15.1% | Corticosteroids | 30.6% | Corticosteroids | 51.8% |
| 6 | H2 receptor antagonist | 28.3% | Corticosteroids | 10.4% | Dexamethasone | 12.1% | ARBs | 8.9% | Aspirin | 29.3% | Ceftriaxone | 39.3% |
| 7 | Famotidine | 27.9% | Vitamin C | 9.7% | Aspirin | 11.8% | Metformin | 6.9% | H2 receptor antagonist | 21.1% | Statins | 32.5% |
| 8 | Aspirin | 26.1% | Metformin | 8.3% | ACE inhibitors | 11.6% | Statins | 4.6% | Famotidine | 20.8% | Aspirin | 27.5% |
| 9 | Alpha-1 blockers | 12.4% | Ceftriaxone | 8.2% | ARBs | 11.5% | Ceftriaxone | 3.6% | Vitamin D | 19.2% | Alpha-1 blockers | 20.8% |
| 10 | ACE inhibitors | 10.9% | Vitamin D | 8.1% | Statins | 10.9% | Dexamethasone | 2.6% | ACE inhibitors | 14.5% | Interleukin inhibitors | 16.5% |

|  | PREMIER |  |  |  | STARR |  | TRDW |  | VA |  |  |  |
| --- | --- | --- | --- | --- | --- | --- | --- | --- | --- | --- | --- | --- |
|  | Hospitalized |  | Intensive Services |  | Hospitalized |  | Hospitalized |  | Hospitalized |  | Intensive Services |  |
| N | Treatment | % | Treatment | % | Treatment | % | Treatment | % | Treatment | % | Treatment | % |
| 1 | Enoxaparin | 49.4% | Enoxaparin | 51.1% | Corticosteroids | 66.7% | Enoxaparin | 58.3% | Vitamin D | 95.2% | Vitamin D | 98.5% |
| 2 | Statins | 32.8% | H2 receptor antagonist | 35.2% | Dexamethasone | 54.0% | Heparin | 51.2% | Enoxaparin | 58.8% | Statins | 62.7% |
| 3 | Aspirin | 25.1% | Famotidine | 35.1% | Heparin | 50.7% | Ceftriaxone | 40.5% | Statins | 58.1% | Enoxaparin | 59.5% |
| 4 | Corticosteroids | 19.7% | Corticosteroids | 33.8% | Alpha-1 blockers | 38.3% | Statins | 36.2% | Aspirin | 40.1% | Heparin | 58.1% |
| 5 | H2 receptor antagonist | 19.2% | Statins | 32.7% | Enoxaparin | 32.8% | Corticosteroids | 34.0% | Heparin | 35.5% | Corticosteroids | 52.8% |
| 6 | Famotidine | 19.1% | Aspirin | 26.9% | Aspirin | 28.6% | Aspirin | 23.6% | Corticosteroids | 35.3% | Aspirin | 43.5% |
| 7 | Direct factor Xa inhibitors | 14.9% | Direct factor Xa inhibitors | 15.6% | Statins | 28.4% | Vitamin D | 17.8% | Ceftriaxone | 35.2% | Ceftriaxone | 38.5% |
| 8 | Vitamin C | 13.0% | Alpha-1 blockers | 14.4% | Famotidine | 23.8% | H2 receptor antagonist | 17.2% | ACE inhibitors | 25.7% | H2 receptor antagonist | 31.7% |
| 9 | Apixaban | 13.0% | Vitamin C | 14.1% | H2 receptor antagonist | 23.8% | Famotidine | 17.2% | Direct factor Xa inhibitors | 18.6% | Famotidine | 29.3% |
| 10 | Alpha-1 blockers | 10.1% | Apixaban | 13.7% | Ceftriaxone | 13.3% | Alpha-1 blockers | 16.3% | Metformin | 18.4% | Alpha-1 blockers | 24.7% |

Figure 2 shows the proportion of users of each of the targeted repurposed therapies in the month after hospital admission (circle) and after initiation of intensive services (triangle, where available) per database. Hydroxychloroquine was the most used therapy, but with great variability in use ranging from < 2% in China to 85.4% in Spain. Chloroquine was used in China (11.5%). Umifenovir was the most common treatment in China, dispensed to 78.3% of patients. Azithromycin was the second in frequency of use, at a highest of 56.5% in Spain. Lopinavir/ritonavir were the third most popular treatments, with great heterogeneity in use, ranging from lowest in USA (2.8% in Optum EHR), higher in South Korea (34.9% in HIRA) and highest in Spain (56.8% in HM). Oseltamivir was variably used from 0.4% in HIRA (South Korea) to 13.2% in NFHCRD (China). Other treatments under study (interferon, itraconazole, and ivermectin) were rarely used (<1%) in any of the databases.

**Figure 2.**
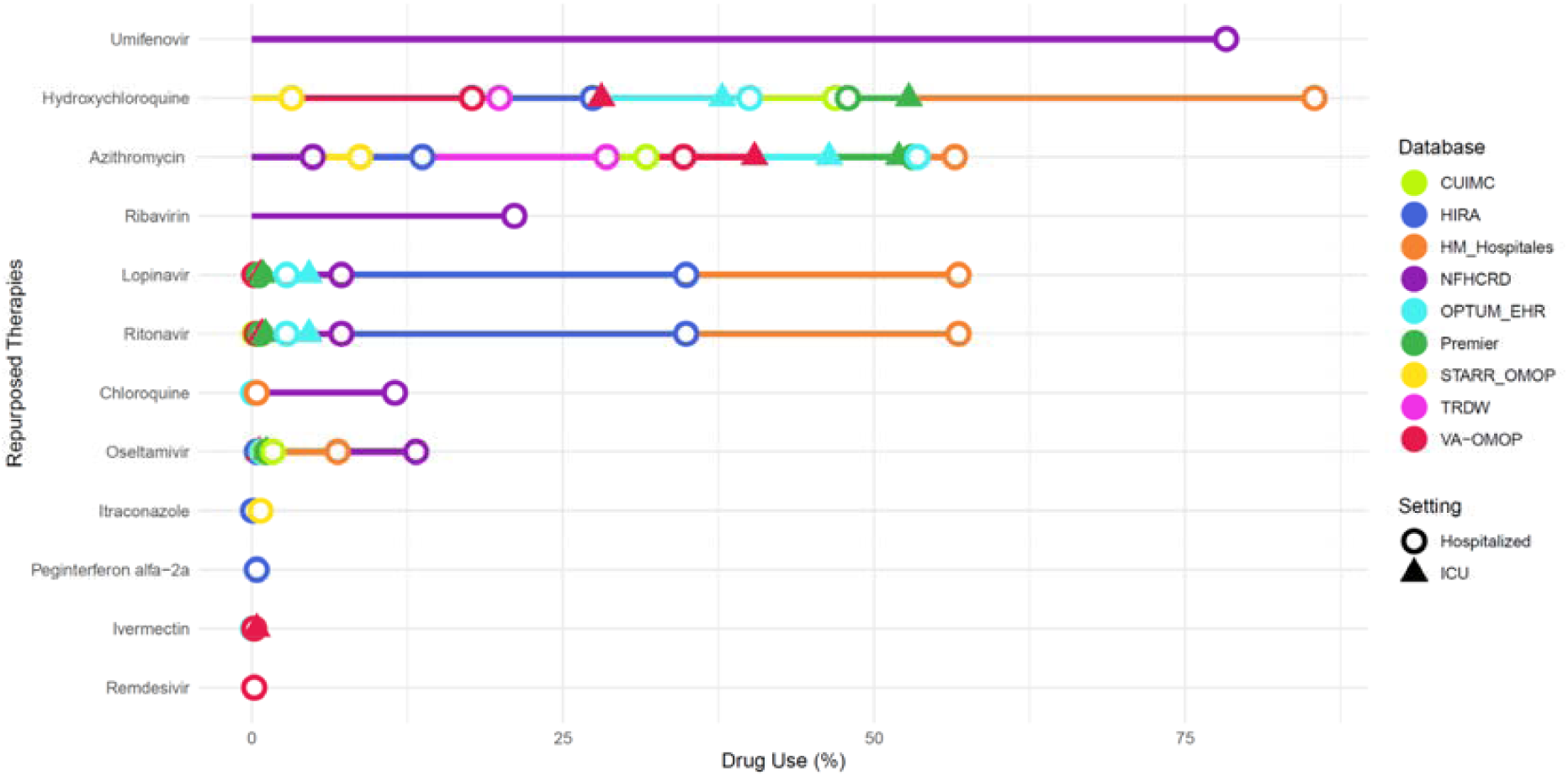
Lollipop plot showing proportion of patients receiving repurposed therapies, in hospitalized or intensive services settings.

The SMD in the proportion of use for each of the listed adjunctive treatments in the month before vs after admission is depicted in Supplementary Figure 1, and the proportion of users in the month after hospital admission for all in Supplementary Figure 2. The list of ‘shortlisted’ therapies with an SMD>0.1 and their respective proportion of users per database is plotted in Figure 3. The ten most used medicines (ingredient level) included, in descending order: bemiparin, enoxaparin, heparin, ceftriaxone, aspirin, vitamin D, famotidine, vitamin C, dexamethasone, and metformin. There was great variability in the use of all these medicines internationally across the participating databases. A 66.7% of people admitted in STARR-OMOP received corticosteroids, compared to lowest proportions in South Korea (10.4%), a 42.8% in Spain, and a 40.8% in China. Statins were also commonly used in the USA (up to 58.1% in VA), but less so in Asia (4.6% in China and 13.5% in South Korea) and Spain (10.9% in HM). Interestingly, vitamin D supplementation was prescribed with great heterogeneity, ranging from <1.6% in China, 1.4% in Spain, 8.1% in South Korea, and a variable 3.4% (Premier) to 95.2% (VA) in the USA. Tocilizumab was also variably used, ranging from <1% in many sites (China, South Korea, NY USA, Premier USA, CA USA) to 4% in VA, 7.0% in Optum EHR and 9.2% in MA USA, and a striking 16.3% in HM (Spain).

**Figure 3.**
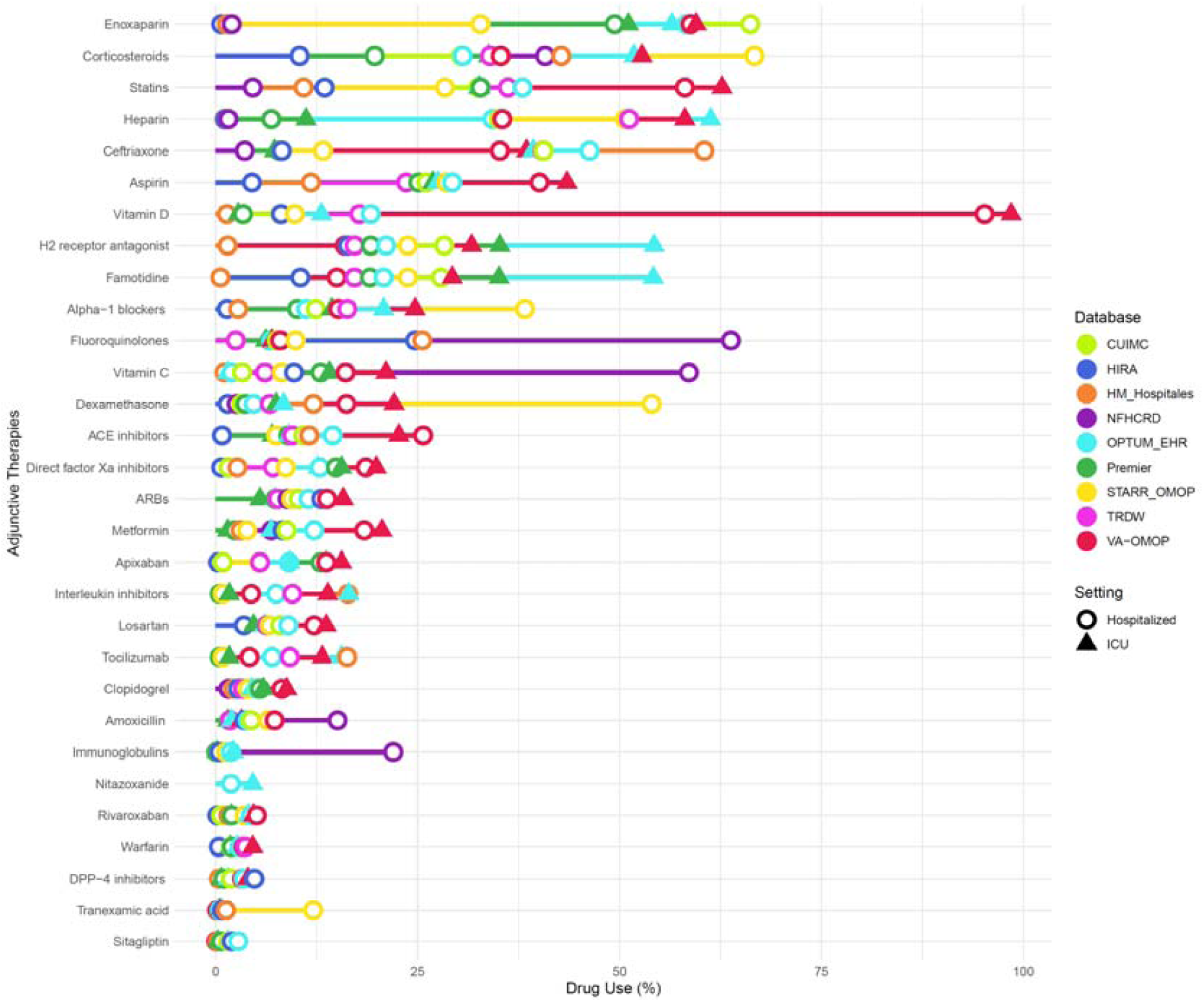
Lollipop plot showing proportion of patients receiving “shortlisted” adjunctive therapies in hospitalized or intensive services settings.

The use of adjunctive therapies (but not of repurposed treatments) increased substantially in intensive care, with the greatest augmentation seen for systemic corticosteroids, famotidine, heparin, and tocilizumab.

The management of COVID-19 has changed substantially over time as shown in Supplementary Figure 3. The time trends in use of hydroxychloroquine are striking, with rapidly increasing use in February and March, a plateau in April, and a similarly rapid decline in May that continued in June (Figure 4). Azithromycin followed a similar trend. Corticosteroids increased in some but not all databases in Apr-May-June.

**Figure 4.**
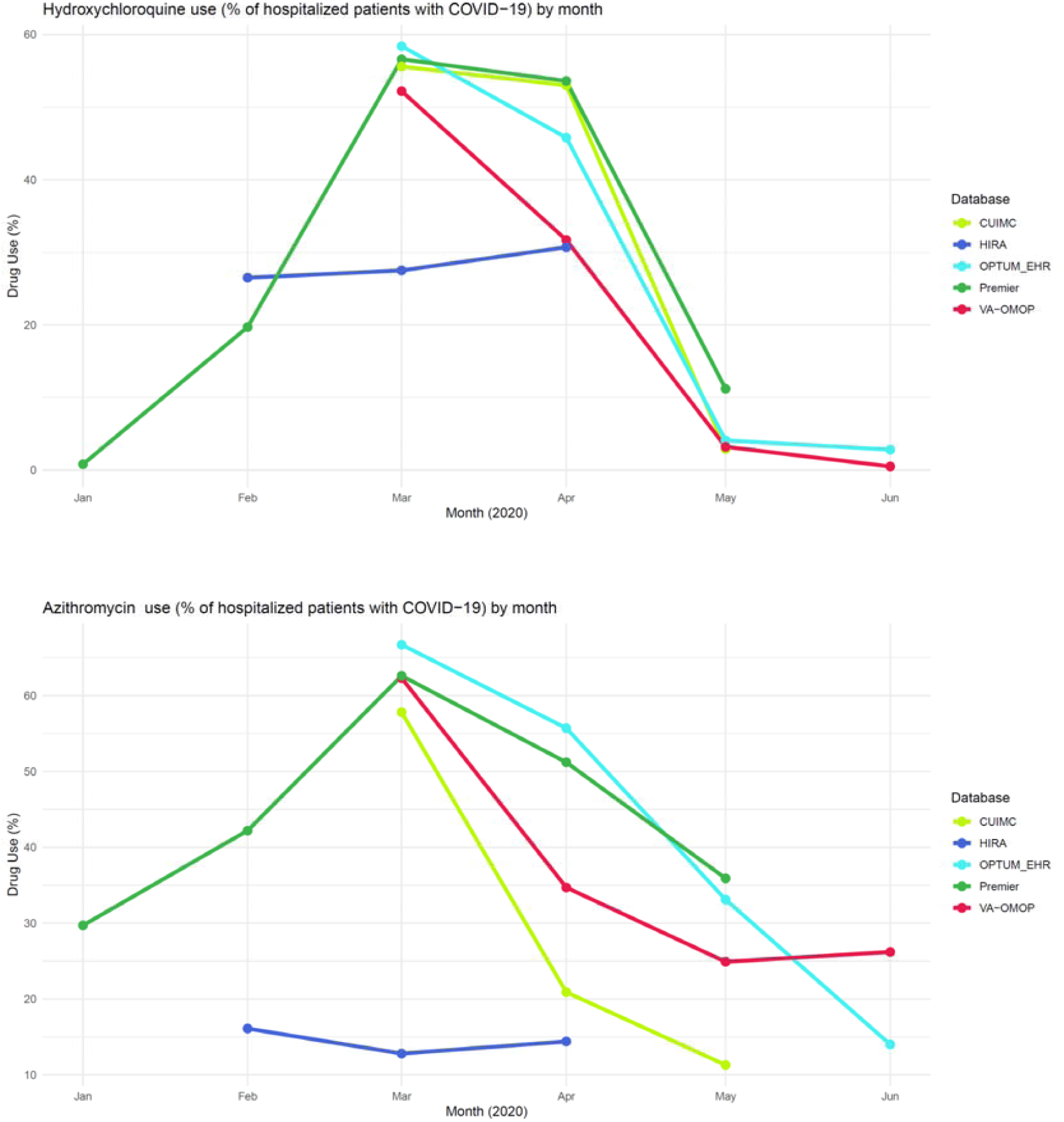
Time trends of hydroxychloroquine and azithromycin in hospitalized patients Jan-June 2020.

## Discussion

### Summary of key findings

This is the first study to describe the management of COVID-19 patients in routine hospital care across three continents. We report on the use of adjunctive and repurposed therapies as recorded in electronic medical records and claims data covering a total of 71,921 hospitalized, and 11,512 patients receiving intensive care for COVID-19 in South Korea, Spain, China and USA.

We observed great heterogeneity in the use of repurposed therapies, with great variability in the use of hydroxychloroquine internationally and over time. Similar trends were observed in the use of azithromycin. Great heterogeneity was also seen in the use of anti-retrovirals, with use of lopinavir/ritonavir ranging from <1% in VA (USA), almost 37% in South Korea, and highest at >50% in Spain.

Adjunctive treatments have been extensively used for the prevention or treatment of complications in the management of COVID19, including antibiotics, anticoagulants, corticosteroids, vitamin D supplements, and to a lesser degree, antihypertensives, antacids, statins and metformin. Unsurprisingly, the use of adjunctive therapies increased amongst patients receiving intensive care services.

Hydroxychloroquine has been in the public limelight since the start of the pandemic. Its use has been supported/endorsed by misleading evidence due to flawed but heavily publicized studies ^12-14^. Despite all the hope and hype, numerous RCTs have shown no benefit. The RECOVERY RCT of 1542 hospitalized participants treated with hydroxychloroquine showed no effects on 28-day mortality when compared to usual care ^15^. Another RCT studied the efficacy of hydroxychloroquine as post-exposure prophylaxis in 821 asymptomatic participants, for whom the drug was not shown to prevent any COVID-19 illness after high or moderate-risk exposure to COVID-19 ^11^.

Azithromycin, a macrolide antibiotic with alleged antiviral efficacy against COVID-19, was also widely prescribed in our data. While several guidelines recommend the use of empirical antimicrobial treatment, not all advocate its use ^7^. Currently, there are 47 RCTs ongoing worldwide, which will hopefully shed some light on the efficacy of azithromycin to treat COVID-19. Use of protease inhibitors (PIs) lopinavir-ritonavir was high in South Korea and Spain, with a much lower use in all other databases. This is consistent with Korean and Spanish guidelines, which recommended PIs as antiviral treatments ^16,17^, probably based on *in-vitro* studies ^18^. The World Health Organization has recently decided to discontinue hydroxychloroquine and lopinavir/ritonavir arms in its SOLIDARITY trial, due to interim results demonstrating little or no reduction in mortality of hospitalized patients ^19^. The RECOVERY trial has recently confirmed the lack of efficacy of lopinavir/ritonavir compared to usual care ^20^. Umifenovir in China was the most prescribed repurposed medicine, consistent with Chinese guidelines and research.^21,22^

Adjunctive therapy/ies to prevent or treat complications were strikingly different across the globe. Heparin use was widely prescribed across the USA and Spain, but not in China and South Korea. Corticosteroids use ranged from about 10% of admitted participants in HIRA (South Korea) to more than 66% of patients in Stanford (CA, USA). Antibiotics use also varied widely, as did statins.

Use of anticoagulants in our study was higher than expected. Severe COVID-19 has been associated with a coagulopathy that, when untreated, leads to poor clinical outcomes ^23^. While a number of RCTs are ongoing to evaluate the value of anticoagulation in patients with COVID-19, interim guidelines recommend the use of anticoagulants for the prophylaxis of thromboembolism ^17,24^. Prior to the results from the RECOVERY trial, there was a wide debate on whether corticosteroids have a role in the mitigation of inflammatory organ injury ^25,26^. Most clinical guidelines did not recommend the use of corticosteroids in COVID-19 ^7^, with notable exceptions ^12,27^. Our results show wide use of corticosteroids internationally.

Our description of patterns in hydroxychloroquine use before and after the publication of negative RCT results indicated a reversal of pre-existing trends. The drastic decline in prescribing pattern is an indication of how rapidly the landscape of medication use changes with the emergence of new evidences. The reverse is also possible, - use of dexamethasone in patients receiving invasive mechanical ventilation or oxygen therapy would likely increase exponentially in-line with recent results from the RECOVERY trial ^5^.

In an attempt to add context to our findings, we conducted a literature review of articles that reported medication use in patients with COVID-19, of which 45% (n=595) reported at least one type of treatment for hospitalized patients. The most reported repurposed therapies were antivirals (25%), antibiotics (19%) and steroids (15%). It is worth noting though that 52% of the studies were from China, whereas our data sources were mainly from USA, Spain and South Korea. In addition, medication use has changed rapidly with the emergence of new evidences, thus findings from China may no longer be relevant for comparison as the studies were older.

### Study limitations

Our study was based on routinely collected real world (EHR and claims) data, where misclassification of disease and therapies may be present. We only included patients who had a clinical COVID-19 diagnosis or a positive PCR test at the time of hospitalization; therefore, patients without a coded diagnosis would have been excluded even if they were suspected of having the disease. There may also be an underreporting of COVID-19 cases in clinical settings where testing resources were scarce, especially during the peak of the outbreak. In addition, medical conditions may be underreported as the absence of a medical code for the disease is interpreted as an absence of the disease itself. Exposure misclassification is also possible; participating data sources varied in their capture of drugs, from hospital billing records, prescription orders, or dispensing data. Medication use estimates on the date of hospitalization is particularly sensitive to misclassification and may conflate baseline concomitant drug history with immediate treatment upon admission. Another limitation was the inability to assess the prescribing pattern of remdesivir as data was not available in our study. The lack of information on the dose and duration of medications was another limitation as these are important information that would have added value to the understanding of prescribing trends, especially among those in high-risk groups or those who are more susceptible to medication-related adverse events. While our study adds valuable information to the understanding of prescribing patterns at the peak of the outbreak, it only provides a snapshot of medication use in clinical practice, and with the constant emergence of new evidences over time, medication use in COVID-19 is likely to evolve rapidly.

### Conclusions

This is the largest and most diverse study characterizing the management of patients hospitalized with COVID-19, covering the first 6 months of the pandemic and spanning across North America, Europe, and Asia. There has been great interest in the safety and efficacy of medications used for COVID-19 treatment, but little evidence on the prescribing patterns in routine clinical practice. This study provides an overview of drug utilization in routine practice and highlights the need for future research on the safety and efficacy of the more commonly used treatments.

## Data Availability

Analyses were performed locally, and the data is not readily available to be shared. However, all analytic code is available at: https://github.com/ohdsi-studies/Covid19CharacterizationCharybdis and results are available at https://data.ohdsi.org/Covid19CharacterizationCharybdis/

https://github.com/ohdsi-studies/Covid19CharacterizationCharybdis

## Competing interest statement

All authors have completed the ICMJE uniform disclosure form at www.icmje.org/coi_disclosure.pdf and declare: Dr Prieto-Alhambra reports grants and other from AMGEN; grants, non-financial support and other from UCB Biopharma; grants from Les Laboratoires Servier, outside the submitted work; and Janssen, on behalf of IMI-funded EHDEN and EMIF consortiums, and Synapse Management Partners have supported training programs organized by DPA’s department and open for external participants. : SCY reports grants from Korean Ministry of Health & Welfare, grants from Korean Ministry of Trade, Industry & Energy, during the conduct of the study; AS reports personal fees from Janssen Research & Development, during the conduct of the study; personal fees from Janssen Research & Development, outside the submitted work; KK is an employee of IQVIA; HAb is an employee and shareholder of Eli Lilly and Company; AA is an employee at Alberta Health Services (AHS) and has been redeployed to aid in the COVID-19 response, this work was not conducted at AHS, within AHS working hours, or with AHS staff, but as a member of the Observational Health Data Sciences and Informatics (OHDSI) Network; JC reports grants from Korean Ministry of Health & Welfare, grants from Korean Ministry of Trade, Industry & Energy, during the conduct of the study; FJDF reports personal fees from Janssen Research & Development, during the conduct of the study; personal fees from Janssen Research & Development, outside the submitted work; WURA reports funding from the NIHR Oxford Biomedical Research Centre (BRC), Aziz Foundation, Wolfson Foundation, and the Royal College Surgeons of England; SLDV reports grants from Anolinx, LLC, grants from Astellas Pharma, Inc, grants from AstraZeneca Pharmaceuticals LP, grants from Boehringer Ingelheim International GmbH, grants from Celgene Corporation, grants from Eli Lilly and Company, grants from Genentech Inc., grants from Genomic Health, Inc., grants from Gilead Sciences Inc., grants from GlaxoSmithKline PLC, grants from Innocrin Pharmaceuticals Inc., grants from Janssen Pharmaceuticals, Inc., grants from Kantar Health, grants from Myriad Genetic Laboratories, Inc., grants from Novartis International AG, grants from Parexel International Corporation through the University of Utah or Western Institute for Biomedical Research outside the submitted work; WG is an AbbVie employee; AG reports personal fees from Regeneron Pharmaceuticals, and full-time employment at Regeneron Pharmaceuticals, outside the submitted work; JH is a full-time employee of Janssen Research & Development, and a shareholder of Johnson & Johnson; GH reports grants from US NIH National Library of Medicine, during the conduct of the study; grants from Janssen Research, outside the submitted work; HJ reports grants from Korean Ministry of Health & Welfare, grants from Korean Ministry of Trade, Industry & Energy, during the conduct of the study; YJ is an AbbVie employee and shareholder; BSKH reports grants from Innovation Fund Denmark (5153-00002B) and the Novo Nordisk Foundation (NNF14CC0001), outside the submitted work; RM is an employee of Janssen Research and Development; DRM reports funding support from the Wellcome Trust, NIHR, Scottish CSO and Tenovus Scotland for research unrelated to this work; RWP reports grants from Korean Ministry of Health & Welfare, grants from Korean Ministry of Trade, Industry & Energy, during the conduct of the study; JP reports grants from Korean Ministry of Health & Welfare, grants from Korean Ministry of Trade, Industry & Energy, during the conduct of the study; APU reports grants from Fundacion Alfonso Martin Escudero, grants from Medical Research Council, outside the submitted work; GAR is a full time employee of Janssen and shareholder of Johnson & Johnson; CR is an employee of IQVIA; PR reports grants from Innovative Medicines Innitiative, grants from Janssen Research and Development, during the conduct of the study; MS is a full-time employee of Janssen R&D, and a shareholder of Johnson & Johnson; AS is a full time employee of Janssen and shareholder of Johnson & Johnson; MAS reports grants from US National Science Foundation, grants from US National Institutes of Health, grants from IQVIA, personal fees from Janssen Research and Development, during the conduct of the study; JS was a full-time employees of Johnson & Johnson, or a subsidiary, at the time the study was conducted, and owns stock, stock options, and pension rights from the company; DV reports personal fees from Bayer, outside the submitted work, and full-time employment at Bayer; DPA reports grants and other from AMGEN, grants, non-financial support and other from UCB Biopharma, grants from Les Laboratoires Servier, outside the submitted work; and Janssen, on behalf of IMI-funded EHDEN and EMIF consortiums, and Synapse Management Partners have supported training programs organized by DPA’s department and open for external participants; PR is an employee of Janssen Research and Development and shareholder of Johnson & Johnson; VS reports funding from US National Science Foundation, funding from the Agency for Healthcare Research and Quality through the University of Utah, and funding from the Arizona Board of Regents. FN was an employee of AstraZeneca until 2019, before the conduct of this study, and owns some AstraZeneca shares. LYHL, EB, DD, MTFA, HAl, OA, TMA, CA, JMB, ACC, AD, TDS, TF, VH, CYJ, DK, SK, YK, SK, JL, HL, KEL, MEM, PM, KN, AO, JDP, YR, LMS, NHS, SS, MS, SV, HW, AEW, BBY, LZ, JJ, MR, ER, HZ, MG and OZ have nothing to report. No other relationships or activities that could appear to have influenced the submitted work.

## Transparency declaration

Lead authors affirm that the manuscript is an honest, accurate, and transparent account of the study being reported; that no important aspects of the study have been omitted; and that any discrepancies from the study as planned have been explained.

## Ethical approval

All the data partners received Institutional Review Board (IRB) approval or exemption. STARR-OMOP had approval from IRB Panel #8 (RB-53248) registered to Leland Stanford Junior University under the Stanford Human Research Protection Program (HRPP). The use of VA data was reviewed by the Department of Veterans Affairs Central Institutional Review Board (IRB) and was determined to meet the criteria for exemption under Exemption Category 4(3) and approved the request for Waiver of HIPAA Authorization. The research was approved by the Columbia University Institutional Review Board as an OHDSI network study. The IRB number for use of HIRA data was AJIB-MED-EXP-20-065). The use of HM Hospitals was approved by the Clinical Research Ethics Committee of the IDIAPJGol (project code: 20/070-PCV). The collection and usage of the data for clinical research in NFHCRD was approved by the IRB of Nanfang Hospital.

## Funding and study sponsors

The European Health Data & Evidence Network has received funding from the Innovative Medicines Initiative 2 Joint Undertaking (JU) under grant agreement No 806968. The JU receives support from the European Union’s Horizon 2020 research and innovation programme and EFPIA. This research received partial support from the National Institute for Health Research (NIHR) Oxford Biomedical Research Centre (BRC), US National Institutes of Health, US Department of Veterans Affairs, Janssen Research & Development, and IQVIA. This work was also supported by the Bio Industrial Strategic Technology Development Program (20001234) funded by the Ministry of Trade, Industry & Energy (MOTIE, Korea) and a grant from the Korea Health Technology R&D Project through the Korea Health Industry Development Institute (KHIDI), funded by the Ministry of Health & Welfare, Republic of Korea [grant number: HI16C0992]. This study was supported by National Key Research & Development Program of China (Project No. 2018YFC0116901). Personal funding included Versus Arthritis [21605], Medical Research Council Doctoral Training Partnership (MRC-DTP) [MR/K501256/1] (JL); MRC-DTP [MR/K501256/1, MR/N013468/1] and Fundación Alfonso Martín Escudero (FAME) (APU); Innovation Fund Denmark (5153-00002B) and the Novo Nordisk Foundation (NNF14CC0001) (BSKH); VINCI [VA HSR RES 13-457] (SLD, MEM, KEL); and NIHR Senior Research Fellowship (SRF-2018-11-ST2-004, DPA). The University of Oxford received funding related to this work from the Bill & Melinda Gates Foundation (Investment ID INV-016201 and INV-019257). No funders had a direct role in this study. The views and opinions expressed are those of the authors and do not necessarily reflect those of the Clinician Scientist Award programme, NIHR, Department of Veterans Affairs or the United States Government, the Ministry of Science and Technology of China, NHS or the Department of Health, England.

## Acknowledgements

The authors appreciate the Korean Health Insurance Review and Assessment Service for providing the data and HM Hospitals for making their data publicly available as part of the COVID Data Save Lives project.

## Contributorship statement

All authors have completed the ICMJE uniform disclosure form at www.icmje.org/coi_disclosure.pdf and declare

## Notes

### Summary of Updates

Author list had a typo in one author (Kristin Kosta should have been Kristin Kostka) and added one author that we forgot first time Andrew Williams

